# Development of polyphenotypic scores to prioritize detection of *G6PD* rs1050828-T carriers in African and African American populations

**DOI:** 10.1101/2025.07.17.25331703

**Authors:** Tianyuan Lu, David Stein, Wenmin Zhang, Yuval Itan, Andrew D. Paterson

**Affiliations:** Department of Population Health Sciences, School of Medicine and Public Health, University of Wisconsin-Madison, Madison, WI, USA; Department of Biostatistics and Medical Informatics, School of Medicine and Public Health, University of Wisconsin-Madison, Madison, WI, USA; Center for Precision Medicine, University of Wisconsin-Madison, Madison, WI, USA; Center for Genomic Science Innovation, University of Wisconsin-Madison, Madison, WI, USA; Center for Demography of Health and Aging, University of Wisconsin-Madison, Madison, WI, USA; Department of Genetics and Genomic Sciences, Icahn School of Medicine at Mount Sinai, New York, NY, USA; Department of Statistics, Faculty of Science, University of Manitoba, Winnipeg, MB, Canada; The Charles Bronfman Institute for Personalized Medicine, Icahn School of Medicine at Mount Sinai, New York, NY, USA; Mindich Child Health and Development Institute, Icahn School of Medicine at Mount Sinai, New York, NY, USA; Genetics and Genome Biology, The Hospital for Sick Children, Toronto, ON, Canada; Division of Epidemiology, Dalla Lana School of Public Health, University of Toronto, Toronto, ON, Canada; Division of Biostatistics, Dalla Lana School of Public Health, University of Toronto, Toronto, ON, Canada

## Abstract

**Introduction:** The *G6PD* missense variant rs1050828-T is common in African and African American populations. It lowers HbA1c levels independent of glycemia, increasing the risk of underdiagnosis or delayed diagnosis of diabetes. We aimed to develop polyphenotypic scores (PPS) using routinely collected phenotypes to identify likely carriers.

**Materials and Methods:** Using data from 31,083 African or African American participants in the All of Us Research Program (AoU), we developed sex-specific PPS through multi-stage variable selection. We validated the PPS in independent African or African American individuals from AoU (N = 8,846), BioMe Biobank (N = 8,839), and UK Biobank (N = 6,811), and evaluated their utility among individuals without diagnosed diabetes.

**Results:** The PPS achieved an area under the receiver operating characteristic curve ranging from 0.7614 to 0.8686 in males and from 0.6033 to 0.6933 in females for identifying rs1050828-T carriers. Among individuals without diagnosed diabetes and with HbA1c <6.5%, PPS-based screening reduced the number needed to screen for identifying one carrier by 65-90% in males and 40-60% in females, compared to random screening.

**Discussion:** The PPS may help identify likely rs1050828-T carriers in African and African American populations, supporting prioritization for targeted genetic testing or alternative diagnostic approaches for diabetes.

## Introduction

Glucose-6-phosphate dehydrogenase (G6PD) deficiency is one of the most common enzyme deficiencies globally, which is estimated to affect around 400 million people^1,2^. G6PD has an important role in the pentose phosphate pathway, which is essential for maintaining redox balance and protecting erythrocytes from oxidative damage^3,4^. The *G6PD* missense variant rs1050828-T (Human Genome Variation Society nomenclature: NP_000393.4:p.Val98Met, NP_001346945.1:p.Val68Met, or NP_001035810.1:p.Val68Met; GRCh38 chrX:154,536,002),located in the non-pseudoautosomal region on the X chromosome, is a well-characterized genetic cause of G6PD deficiency^5,6^. This variant can lead to reduced G6PD enzymatic activity, which directly impairs erythrocyte metabolism by compromising the ability of erythrocytes to detoxify reactive oxygen species and increasing the risk of hemolysis under stressors^1,2,7^.

The rs1050828-T allele is common among individuals of African and African American ancestries^8,9^, with an average allele frequency of approximately 12% in gnomAD reference populations (version 4.1.0), and rare in populations of other ancestries with an allele frequency < 1%^10^. However, its frequency has been shown to vary widely across different populations in sub-Saharan Africa^11^.

The variant has been robustly associated with several blood cell indices and biochemical traits in large genome-wide association studies^12–16^. One notable and clinically meaningful association is between rs1050828 and glycated hemoglobin (HbA1c) levels^12–14^. HbA1c is a widely used biomarker that reflects the average blood glucose over the preceding 2-3 months and serves as one of the primary tools for diagnosing and monitoring diabetes^17^. For example, a diagnosis of diabetes can be made with an HbA1c level ≥ 6.5% (48 mmol/mol) based on the American Diabetes Association guidelines^18^. However, the rs1050828-T allele lowers HbA1c levels through non-glycemic mechanisms, primarily by shortening erythrocyte lifespan, thereby reducing the time available for hemoglobin to undergo glycation^12^. Consequently, individuals carrying this variant may present with HbA1c levels below the diagnostic threshold for diabetes despite having hyperglycemia, leading to potential underdiagnosis or delayed diagnosis of diabetes^12,19,20^. Several studies have proposed that HbA1c-based diagnostic thresholds for diabetes should be adjusted in individuals with G6PD deficiency to avoid clinical misclassification. For example, male hemizygotes with an HbA1c level ≥ 5.7% (39 mmol/mol), female homozygotes with a level ≥ 5.8% (40 mmol/mol), and female heterozygotes with a level ≥ 6.2% (44 mmol/mol) may meet diagnostic criteria for diabetes^12^.

Despite its clinical relevance, routine screening for rs1050828-T has not been widely implemented, largely due to constraints in genotyping capacity and cost-effectiveness. However, with the rapid reduction in genotyping or sequencing costs and continuous efforts to integrate genomics into healthcare, targeted screening is becoming more feasible^21^. Nevertheless,genotyping or sequencing the entire population remains impractical in many settings, which necessitates efficient strategies to identify individuals who are most likely to carry the variant and may benefit most from confirmatory genetic testing or alternative diagnostic approaches for diabetes, such as fasting glucose test^18^.

Given that the variant demonstrates measurable effects on inexpensive, routinely collected hematological and biochemical phenotypes that are widely available in electronic health records, we hypothesized that polyphenotypic scores (PPS) aggregating multiple phenotype measurements could be used to predict rs1050828-T carrier status. We developed and optimized sex-specific PPS using data from African or African American participants in the All of Us Research Program (AoU; N = 31,083)^22^. We evaluated the performance of the PPS in identifying rs1050828-T carriers across three test cohorts, including an independent subset of the AoU (N = 8,846), as well as the Mount Sinai BioMe Biobank (BioMe; N = 8,839)^23^, and the UK Biobank (UKB; N = 6,811)^24^. Finally, we assessed the potential clinical utility of the PPS in prioritizing individuals without a known diagnosis of diabetes for confirmatory genetic testing. This study may provide a new strategy for improving G6PD deficiency screening, as well as new opportunities to enhance the prevention and diagnosis of diabetes.

## Methods

### The All of Us Research Program

We leveraged the AoU (version 8) to develop sex-specific PPS for identifying individuals likely to carry the rs1050828-T allele. The AoU began participant enrolment in 2018 and collected data through multiple sources, including electronic health records, self-reported surveys, physical measurements, and biospecimen collection^22^. Whole-genome sequencing was performed on DNA extracted from blood samples using the Illumina NovaSeq 6000 platform, with standardized protocols for quality control and data harmonization. Individuals whose DNA samples failed sample outlier quality control were excluded from analysis. For this study, we included participants who self-identified as “Black or African American”. We used self-reported race rather than genetically inferred ancestry to reflect a practical clinical context in which genetic data are not typically available at the point of care.

We considered 19 phenotypes as candidate predictors for the PPS, including body mass index (BMI), waist-hip ratio (WHR), alanine aminotransferase level (ALT), aspartate aminotransferase level (AST), serum albumin level (ALB), total bilirubin level (TBIL), hemoglobin level (Hb), red blood cell distribution width (RDW), basophil count, eosinophil count, erythrocyte count, leukocyte count, lymphocyte count, monocyte count, neutrophil count, platelet count, mean corpuscular hemoglobin (MCH), mean corpuscular hemoglobin concentration (MCHC), and mean corpuscular volume (MCV). We removed outlier values for each phenotype defined as more than two times the interquartile range below the first quartile or above the third quartile. For individuals with multiple measurements of a phenotype, we excluded any measurements taken before the age of 18 years and retained the most recent value. To reduce bias due to relatedness, we further excluded individuals with a kinship coefficient > 0.1.

### Development of a polyphenotypic score

After filtering, we randomly selected 50% of the AoU participants to form a model selection dataset. Within this dataset, we first regressed each of the 19 candidate phenotypes on sex and age at the time of measurement to obtain residualized phenotypes, which were then standardized to have a zero mean and unit variance. To assess associations with carrier status, we then performed sex-stratified univariable logistic regression using the residualized phenotypes. Carriers were defined as hemizygous males, and females with either one or two copies, of the rs1050828-T allele. Phenotypes that were significantly associated with carrier status after Bonferroni correction (p-value < 0.05 / 19 phenotypes / 2 sex-specific tests = 1.3×10^−3^) were selected for inclusion in the PPS.

Next, we restricted the analysis to individuals with non-missing data for all selected phenotypes. We conducted sex-specific multivariable logistic regression using backward elimination to iteratively remove redundant phenotypes using the Bayesian information criterion (BIC)^25^. The logistic regression coefficients from the optimized model with the lowest BIC were used as weights to construct the PPS for males and females, respectively.

### Evaluation of model performance

We evaluated the performance of the PPS in identifying individuals more likely to be carriers of the rs1050828-T allele across three cohorts. First, we formed an AoU test dataset comprising AoU participants who were not included in the model selection dataset. In addition, we leveraged two external cohorts, from the Mount Sinai BioMe Biobank and the UK Biobank. BioMe is an electronic health record-link cohort based in New York City, initiated in 2007^23^. Genetic data in BioMe were generated using the Regeneron Global Screening Array or the Sema4 Global Diversity Array for genotyping, or through whole-exome sequencing performed on the Illumina HiSeq 2500 platform. Participants were recruited from Mount Sinai primary care clinics (https://icahn.mssm.edu/research/ipm/programs/biome-biobank). No specific inclusion criteria were applied during recruitment. Therefore, the participants are expected to be representative of the population of New York City and its surrounding neighborhoods. UKB is a population-based, multi-center cohort in the United Kingdom that enrolled approximately 500,000 participants aged 40-69 years at recruitment between 2006 and 2010^24^. Genotyping in UKB was conducted using the UK BiLEVE Axiom Array or the UK Biobank Axiom Array. The variant of interest, rs1050828, was directly genotyped on these arrays and did not require imputation. We included participants who self-identified as “African American” in BioMe, and “African”, “Caribbean”, “Black or Black British”, or “Any other Black background” in UKB.

In each cohort, we retained individuals with non-missing data for all phenotypes included in the PPS. In the AoU test dataset and BioMe, we excluded any measurements taken before the age of 18 years and retained the most recent value for each phenotype. In UKB, we used phenotype measurements obtained at the baseline assessment. Within each cohort, we regressed each phenotype on sex and age at the time of measurement to obtain residualized values, which were then standardized to have a zero mean and unit variance. We subsequently calculated the PPS for all individuals. We evaluated the performance of the PPS by computing the area under the receiver operating characteristic curve (AUROC) for identifying rs1050828-T carriers, separately by sex. To assess the added value of the PPS, we compared its performance to that of each individual phenotype considered independently. We performed DeLong’s test to examine whether the difference in model performance was significant^26^.

### Evaluation of clinical utility in screening programs

To further evaluate the potential clinical utility of the PPS in refining diagnostic criteria for diabetes, we assessed the efficiency of the PPS in hypothetical screening programs targeting individuals without a known diagnosis of diabetes. In the AoU test dataset, BioMe, and UKB, we retained individuals whose most recent HbA1c measurement was taken after age 18 and who did not have evidence of diabetes based on electronic health records or self-reported physician-made diagnosis. Evidence of diabetes from electronic health records included International Classification of Diseases version 9 (ICD-9) codes under category 250, ICD-10 codes under category E08-E14, a fasting glucose level ≥ 126 mg/dL, a random glucose level ≥ 200 mg/dL, or an HbA1c level ≥ 6.5% (48 mmol/mol) at any time point^18^. We did not incorporate medication codes due to variability in prescribing practices, incomplete capture of medication data, and the potential for off-label use of antidiabetic medications for conditions unrelated to diabetes.

Within each cohort, at various percentile cutoffs of the PPS (top 5%, 10%, 20%, 30%, 40%, and 50%), we calculated the proportion of carriers of the rs1050828-T allele, and correspondingly, the number needed to screen for identifying one carrier. For example, the number needed to screen corresponding to the top 5% PPS cutoff was calculated as:

Number needed to screen = (Number of individuals in the top 5% of PPS specific to the target population) / (Number of carriers in the top 5% of PPS specific to the target population).

We examined two target populations separately: (1) individuals with an HbA1c level ≥ 5.0% (31 mmol/mol) and < 6.5% (48 mmol/mol), and (2) individuals with an HbA1c level < 6.5% (48 mmol/mol). Evaluation among individuals typically classified as having prediabetes (5.7% ≤ HbA1c level < 6.5%, i.e., 39 mmol/mol ≤ HbA1c level < 48 mmol/mol)^18^ was not feasible due to limited sample sizes in all three test cohorts. A lower number needed to screen indicates higher efficiency of the screening program.

## Results

### Study overview

An overview of the study design and cohort characteristics is presented in **Figure 1**. All cohorts comprised individuals who self-identified as African or African American (**Methods**). The AoU model selection dataset included 13,283 males (mean age = 54.1 years; SD = 14.4 years) and 17,800 females (mean age = 52.3 years; SD = 15.4 years) who had genetic data and at least one measurement for any of the 19 candidate phenotypes after quality control. The rs1050828-T allele frequency was 11.8% in males and 12.3% in females (**Figure 1**). The distributions of age at the most recent measurement of each individual phenotype are presented in **Supplementary Table 1**.

**Figure 1.**
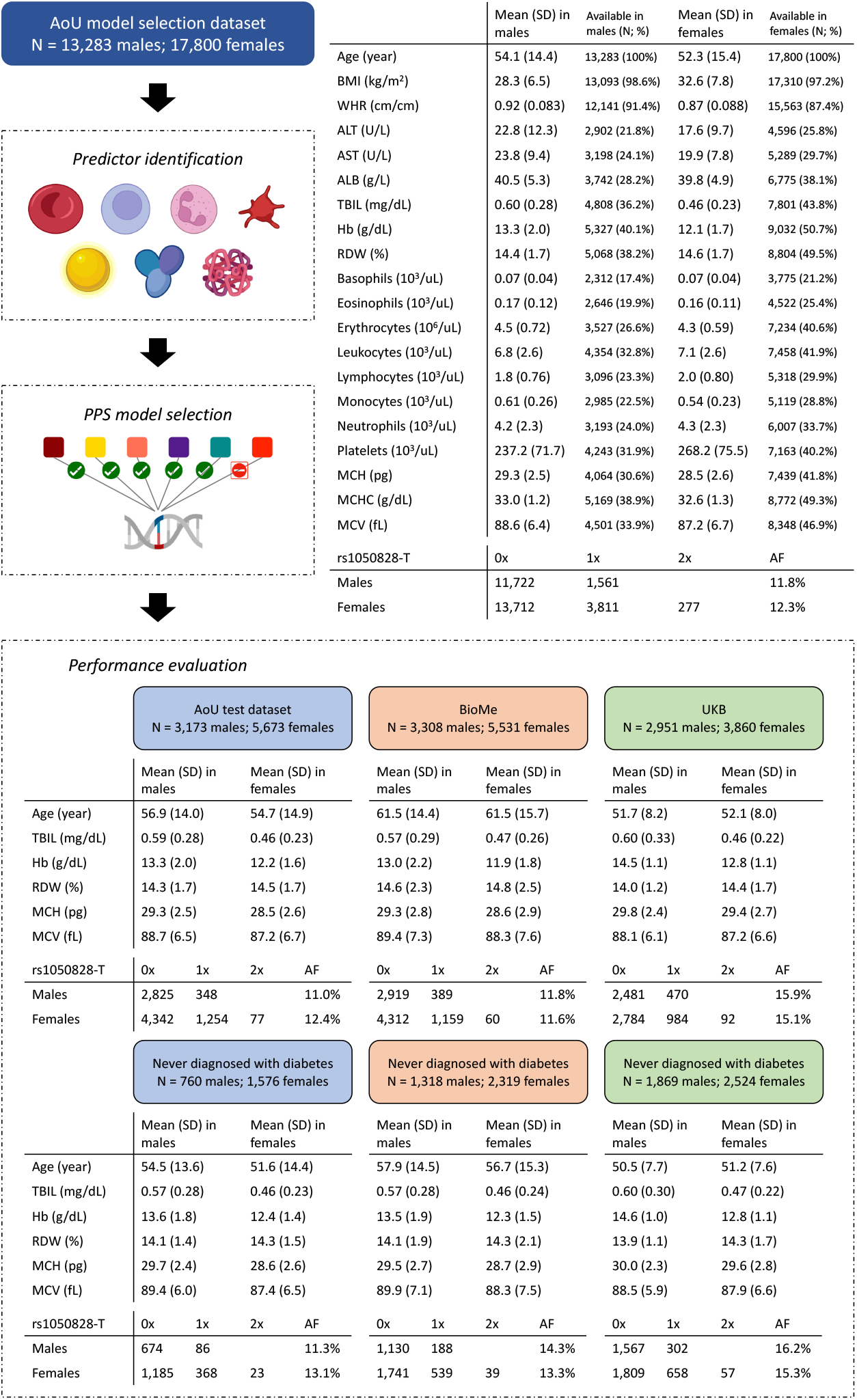
Overview of study design. Sex-specific PPS were developed in the AoU model selection dataset using multi-stage variable selection. Cohort characteristics, including all candidate phenotypes, are summarized for the AoU model selection dataset. The PPS were evaluated in the AoU test dataset, BioMe, and UKB, which do not overlap with the AoU model selection dataset. Cohort characteristics, including selected phenotypes in the PPS, are summarized for these test cohorts, as well as for the respective subsets comprising individuals who had never been diagnosed with diabetes. Ages at the most recent measurement across all phenotypes are summarized.

The performance of the optimized PPS was evaluated in three cohorts with complete data for all phenotypes included in the score. The AoU test dataset included 3,173 males (mean age = 56.9 years; SD = 14.0 years) and 5,673 females (mean age = 54.7 years; SD = 14.9 years). BioMe included 3,308 males (mean age = 61.5 years; SD = 14.4 years) and 5,531 females (mean age = 61.5 years; SD = 15.7 years). UKB included 2,951 males (mean age = 51.7 years; SD = 8.2 years) and 3,860 females (mean age = 52.1 years; SD = 8.0 years). The rs1050828-T allele frequency was similar between the AoU test dataset (11.0% in males; 12.4% in females) and BioMe (11.8% in males; 11.6% in females) and higher in UKB (15.9% in males; 15.1% in females; **Figure 1**). The variant was in Hardy-Weinberg equilibrium among females across all datasets analyzed (**Supplementary Table 2**).

Although phenotype levels may differ between males and females, the mean levels for males and for females, respectively, were consistent across cohorts (**Figure 1**).

### Polyphenotypic score for carrier status

Univariable logistic regression identified six phenotypes that were significantly associated with carrier status of the rs1050828-T allele (Bonferroni-corrected p-value < 0.05; **Figure 2A** and **Supplementary Table 3**) in both males and females. Specifically, in males, a one standard deviation increase in TBIL, Hb, RDW, erythrocyte count, MCH, and MCV was associated with odds ratios of 1.04 (95% CI: 1.03-1.05; p-value = 8.1×10^−21^), 0.98 (95% CI: 0.98-0.99; p-value = 5.8×10^−5^), 0.93 (95% CI: 0.92-0.94; p-value = 4.8×10^−53^), 0.96 (95% CI: 0.95-0.97; p-value = 1.4×10^−16^), 1.05 (95% CI: 1.04-1.06; p-value = 1.2×10^−24^), and 1.07 (95% CI: 1.06-1.08; p-value = 5.6×10^−44^), respectively. In females, the corresponding odds ratios were 1.05 (95% CI: 1.03-1.06; p-value = 4.5×10^−18^), 0.97 (95% CI: 0.96-0.98; p-value = 1.8×10^−8^), 0.96 (95% CI: 0.95-0.97; p-value = 3.2×10^−22^), 0.95 (95% CI: 0.94-0.96; p-value = 2.1×10^−25^), 1.04 (95% CI: 1.03-1.05; p-value = 6.7×10^−13^), and 1.05 (95% CI: 1.04-1.06; p-value = 3.5×10^−28^).

**Figure 2.**
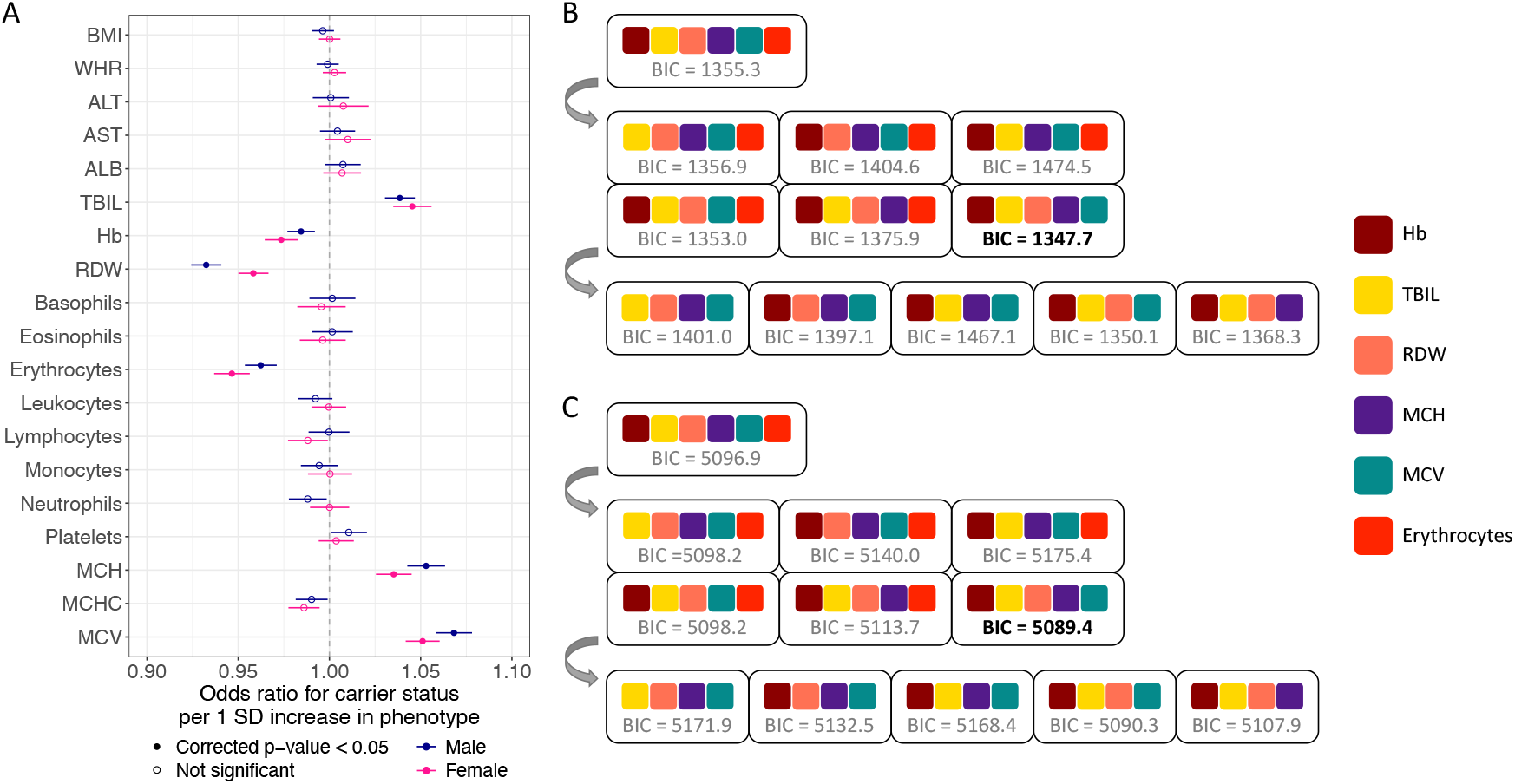
Development of sex-specific PPS in the AoU model selection dataset. (A) Sex-specific marginal associations between candidate phenotypes and rs1050828-T carrier status. Significant associations (Bonferroni-corrected p < 0.05) are shown as solid dots. Non-significant associations are shown as empty dots. Error bars represent 95% confidence intervals. (B) Backward model selection for developing the male-specific PPS. (C) Backward model selection for developing the female-specific PPS. Phenotypes included in each multivariable logistic regression model are represented by colored squares. Final models were selected based on the lowest BIC.

These six phenotypes were used to develop the sex-specific PPS (**Figure 2B and 2C**). Given correlations among these phenotypes (**Supplementary Figure 1**), backward elimination was applied to optimize the final model for each sex based on BIC. As a result, in both males and females, the optimized PPS included TBIL, Hb, RDW, MCH, and MCV as predictors, excluding erythrocyte count.

#### The male-specific PPS was constructed as

PPS = 0.46883 x standardized TBIL + (−0.59085) x standardized Hb + (−1.09429) x standardized RDW + (−0.57592) x standardized MCH + 0.95737 x standardized MCV

#### The female-specific PPS was constructed as

PPS = 0.25770 x standardized TBIL + (−0.42089) x standardized Hb + (−0.40519) x standardized RDW + (−0.26166) x standardized MCH + 0.43053 x standardized MCV

### Model performance in test cohorts

In the AoU test dataset, BioMe, and UKB, the PPS consistently and substantially outperformed each individual phenotype in identifying carriers of the rs1050828-T allele in both sexes (**Figure 3** and **Supplementary Table 4**). Specifically, in the AoU test dataset, the PPS achieved an AUROC of 0.8089 in males and 0.6376 in females. The best-performing individual phenotype was RDW, which yielded an AUROC of 0.7254 in males (DeLong’s test p-value = 4.3×10^−12^) and 0.5929 in females (DeLong’s test p-value = 1.0×10^−7^).

**Figure 3.**
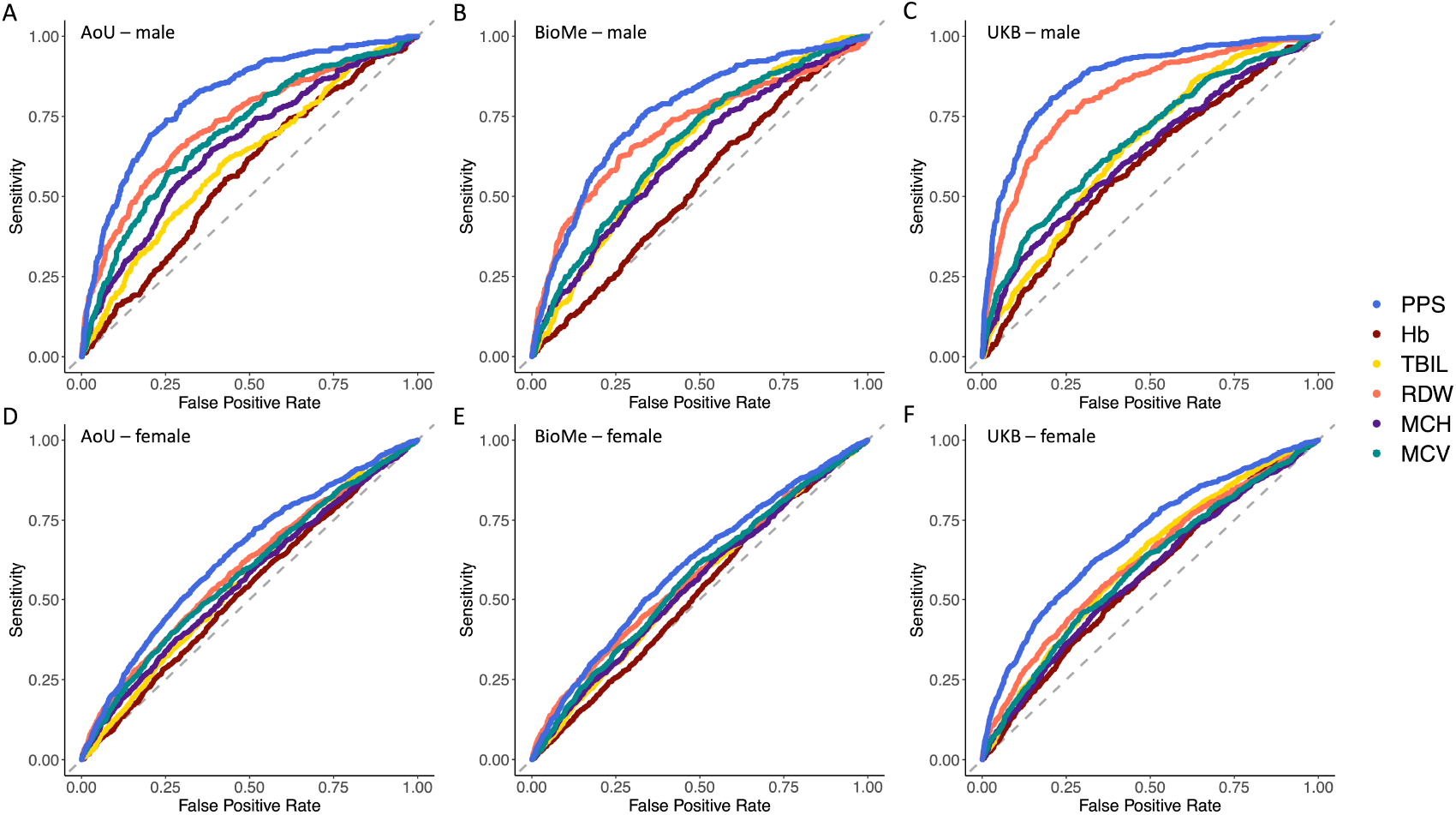
Improved performance of the PPS in identifying rs1050828-T carriers across test cohorts. Sex-specific receiver operating characteristic curves are displayed for (A) males in the AoU test dataset, (B) males in BioMe, (C) males in UKB, (D) females in the AoU test dataset, (E) females in BioMe, and (F) females in UKB. Grey dashed lines indicate the expected performance of a non-informative predictor.

In BioMe, the PPS achieved an AUROC of 0.7614 in males and 0.6033 in females, outperforming RDW which achieved 0.7097 in males (DeLong’s test p-value = 5.7×10^−5^) and 0.5752 in females (DeLong’s test p-value = 3.6×10^−3^). In UKB, the PPS achieved an AUROC of 0.8686 in males and 0.6933 in females, while RDW achieved 0.8147 in males (DeLong’s test p-value = 2.7×10^−9^) and 0.6206 in females (DeLong’s test p-value = 4.3×10^−15^). The AUROC differences PPS and phenotypes other than RDW were more substantial (**Figure 3** and **Supplementary Table 4**).

### Efficiency of polyphenotypic score-guided screening programs

The PPS demonstrated potential clinical utility for stratifying individuals without a known diagnosis of diabetes according to their likelihood of carrying the rs1050828-T allele. Among males with an HbA1c level between 5.0% (31 mmol/mol) and 6.5% (48 mmol/mol), 27.8% in the AoU test dataset, 24.1% in BioMe, and 30.6% in UKB of the individuals in the top 5% of the PPS were carriers (**Supplementary Table 5**). This corresponds to a number needed to screen of 3.6, 4.2, and 3.3, respectively (**Figure 4A Supplementary Table 5**). With more lenient PPS cutoffs, more carriers were captured, while the efficiency of the screening program decreased but remained substantially higher than with random screening. For instance, with a 50% cutoff, the number needed to screen was 9.4 in the AoU test dataset, 6.7 in BioMe, and 14.8 in UKB (**Figure4A** and **Supplementary Table 5**). In contrast, screening all males in this HbA1c range at random required screening 17.5 in the AoU test dataset, 11.6 in BioMe, and 28.2 in UKB to identify one carrier (Figure 4A and Supplementary Table 5). If the target population was all males with an HbA1c level < 6.5% (48 mmol/mol), restricting screening to the top 5% by PPS achieved a number needed to screen of 2.2 in the AoU test dataset, 1.9 in BioMe, and 1.4 in UKB, compared to 8.8, 7.0, and 6.2 with random screening (Figure 4A and Supplementary Table 5).

**Figure 4.**
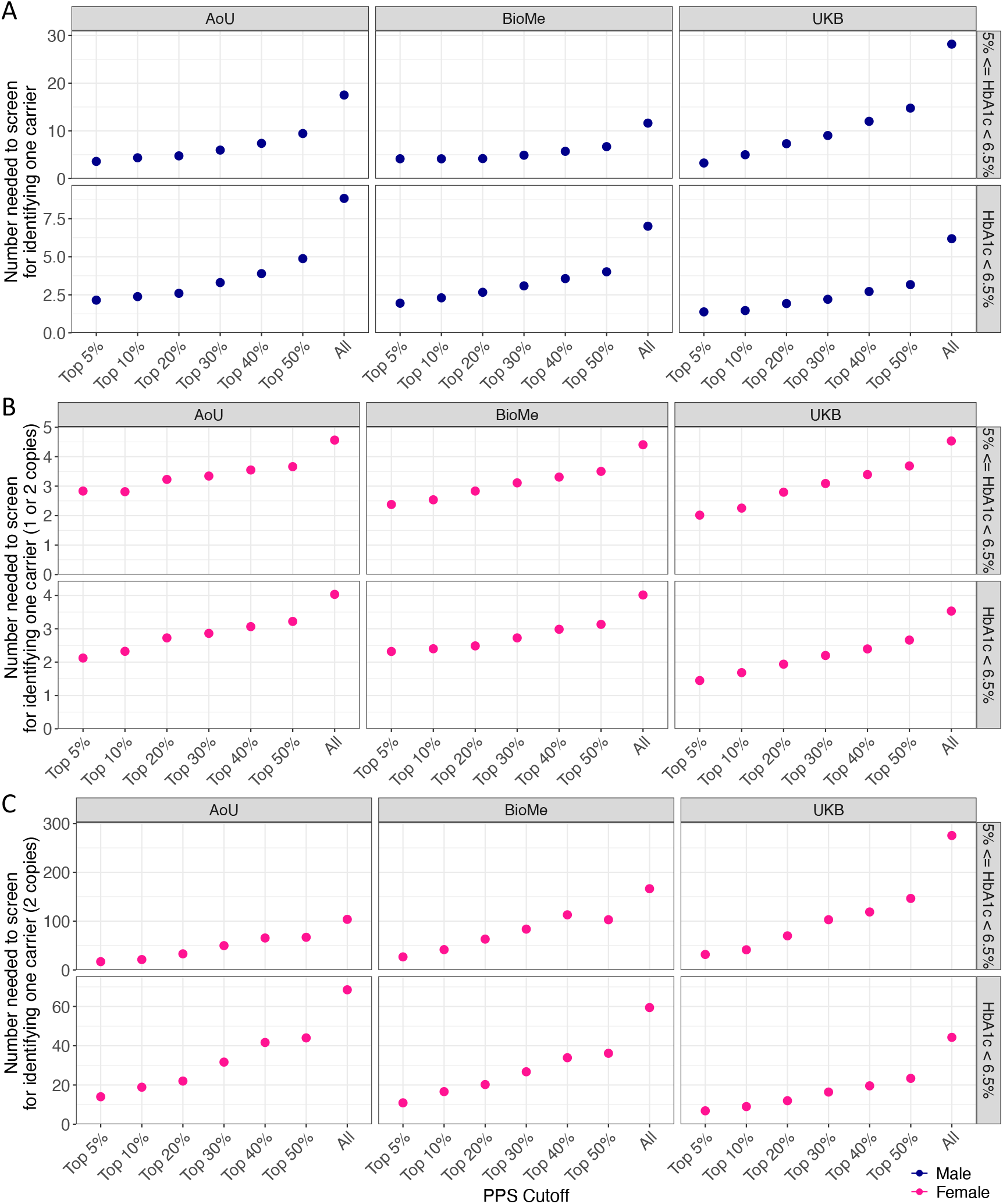
Efficiency of the PPS in hypothetical screening programs targeting individuals without a known diagnosis of diabetes. (A) Number needed to screen for identifying one carrier in males. (B) Number needed to screen for identifying one carrier with at least one copy of the rs1050828-T allele in females. (C) Number needed to screen for identifying one carrier with two copies of the rs1050828-T allele in females. Screening efficiency was assessed separately for individuals with HbA1c levels ≥ 5.0% and < 6.5%, and for those with HbA1c < 6.5%. Values are shown across different PPS cutoffs. When all individuals are included in the target population, the number needed to screen reflects the efficiency of random screening.

Similar patterns were observed in females. For example, among females with an HbA1c level between 5.0% (31 mmol/mol) and 6.5% (48 mmol/mol), the number needed to screen for the top 5% PPS group was 2.8 in the AoU test dataset, 2.4 in BioMe, and 2.0 in UKB, compared to 4.6, 4.4, and 4.5 with random screening (**Figure 4B** and **Supplementary Table 5**). Notably, the PPS more dramatically improved the identification of females with two copies of the rs1050828-T allele. Compared to random screening, the number needed to screen for identifying homozygotes decreased from 103.6 to 17.0 in the AoU test dataset, from 166.4 to 26.8 in BioMe, and from 275.4 to 31.8 in UKB among those in the top 5% of the PPS (**Figure 4C** and **Supplementary Table 5**).

## Discussion

In this study, we developed and validated sex-specific PPS using routinely collected hematologic and biochemical measurements to identify likely carriers of the rs1050828-T allele in African and African American populations. These measurements are widely available in electronic health records, making the approach highly scalable and potentially implementable in real-world clinical settings. We demonstrated that the PPS could meaningfully stratify the population with respect to rs1050828-T carrier status in independent test datasets from three biobanks in the United States and the United Kingdom. These findings suggest that the PPS may be useful for guiding efficient confirmatory genetic testing for G6PD deficiency or alternative diagnostic approaches for diabetes, particularly in settings where genotyping or sequencing is not feasible for all individuals.

The observed associations between the rs1050828-T allele and multiple phenotypes, including TBIL, Hb, RDW, erythrocyte count, MCH, and MCV, are consistent with the known pathophysiology of G6PD deficiency and prior genome-wide association studies^15,16^. Specifically, the rs1050828-T allele impairs G6PD enzyme activity, thus increasing erythrocyte susceptibility to hemolysis^1,2^. This leads to reduced erythrocyte counts and Hb levels, as well as elevated TBIL levels due to increased heme breakdown^27^. Hemolysis and the compensatory erythropoietic response can also influence red cell indices including RDW, MCH, and MCV. These pleiotropic effects support the integration of these phenotypes into a composite score that more accurately captures the biochemical signature of G6PD deficiency.

Our findings have important clinical and translational implications. The rs1050828-T allele is known to lower HbA1c levels through non-glycemic mechanisms, which can complicate diabetes screening and monitoring among affected individuals^12,19,20^. We showed that individuals without a known diagnosis of diabetes but with elevated PPS values were substantially enriched for carriers of the rs1050828-T allele. This provides a pragmatic strategy to prioritize individuals for targeted confirmatory genetic testing or alternative diagnostic approaches for diabetes to mitigate the risk of underdiagnosis or delayed diagnosis. Across the three test cohorts, in males, we observed that screening the top 5% of individuals based on PPS reduced the number needed to screen for identifying one carrier by 65-90% compared to random screening, consistent with the hemizygous expression of X-linked genes. In females, screening the top 5% of individuals based on PPS reduced the number needed to screen by 40-60%, and would decrease further (by 80-90%) if the screening objective were to identify homozygotes, highlighting the utility in various clinical and research scenarios. Notably, the data used for developing the PPS were heterogeneous, with measurements derived from different sources and collected at varying ages across phenotypes. Despite these complexities, the PPS demonstrated robust and consistent performance across cohorts, particularly in UKB, where allele frequency differences may reflect distinct demographic histories. Future investigations may be warranted to explore how similar PPS frameworks can be adapted to other pharmacogenetic or clinically relevant variants, assess their performance in prospective settings, and formally evaluate the cost-effectiveness in clinical implementation.

This study has important limitations. First, the PPS were developed and validated specifically among individuals who self-identified as African or African American, and its performance in other racial, ethnic, or genetic ancestry groups remains unknown. Even within African and African American populations, the performance of the PPS may vary under different environmental, clinical, or healthcare contexts, which have yet to be systematically evaluated. Second, selection bias may be present in the study populations. For example, the notable differences in missing rate and allele frequency between males and females may reflect unknown sources of participation or ascertainment bias^28,29^. Relatedly, the PPS have not been calibrated to yield absolute probabilities of carrier status, as the study populations may not represent the general population. Broader validation and calibration in independent and more diverse cohorts will be valuable. Third, the PPS currently only incorporate the most recent measurements of the phenotypes and does not model longitudinal changes. Incorporating trajectories across multiple visits may further improve its performance, although such efforts must carefully account for age-related trends. Fourth, we were unable to determine the number of rs1050828-T carriers who had been clinically diagnosed with G6PD deficiency, as the available electronic health records did not include diagnostic codes corresponding to the condition. Last, we were unable to specifically isolate individuals in the prediabetic range of HbA1c (5.7-6.0%, i.e., 39-48 mmol/mol) due to limited sample sizes. We instead combined them with individuals having an HbA1c level between 5.0-5.7% (31-39 mmol/mol), which, while not formally considered prediabetes, has been associated with increased risk of future diabetes compared to an HbA1c level < 5.0% (31 mmol/mol)^30–32^. Individuals in this range who are identified as rs1050828-T carriers may also benefit from early preventive interventions such as lifestyle modifications. Future studies may explore the utility of PPS in additional clinical contexts and examine its impact on long-term outcomes.

In conclusion, this study demonstrates that PPS combining routine hematological and biochemical phenotypes may serve as an effective and scalable tool for identifying likely carriers of the rs1050828-T allele in African and African American populations, thereby helping prioritize individuals for confirmatory genetic testing or clinical follow-up. Integrating such scores into clinical workflows may enhance the precision of diabetes diagnosis in high-risk populations and support broader efforts to reduce disparities in genomic medicine.

## Data Availability

Access to the All of Us Research Program is governed by the data access policies of the program and is available to registered researchers in accordance with those guidelines. The UK Biobank and Mount Sinai BioMe Biobank data used in this study are available upon approval by the respective research committees.

## Acknowledgements

We gratefully acknowledge All of Us participants for their contributions, without whom this research would not have been possible. We also thank the National Institutes of Health’s All of Us Research Program for making available the participant data examined in this study. This research has been conducted using the UK Biobank Resource under Application Number 48873. UK Biobank is generously supported by its founding funders the Wellcome Trust and UK Medical Research Council, as well as the Department of Health, Scottish Government, the Northwest Regional Development Agency, British Heart Foundation and Cancer Research UK. The organization has over 150 dedicated members of staff, based in multiple locations across the UK. The Mount Sinai BioMe Biobank has been supported by The Andrea and Charles Bronfman Philanthropies and in part by federal funds from the NHLBI and NHGRI. We thank all participants in the Mount Sinai Biobank. We also thank all our recruiters who have assisted and continue to assist in data collection and management and are grateful for the computational resources and staff expertise provided by Scientific Computing at the Icahn School of Medicine at Mount Sinai. T.L. has been supported by start-up funding from the Office of the Vice Chancellor for Research and Graduate Education, School of Medicine and Public Health, and Department of Population Health Sciences at the University of Wisconsin-Madison.

## Data Availability

Access to the All of Us Research Program is governed by the program’s data access policies and is available to registered researchers in accordance with those guidelines. The UK Biobank and Mount Sinai BioMe Biobank data used in this study are available upon approval by the respective research committees.

## Author Contributions

T.L. and A.D.P. conceptualized the study. T.L. and W.Z. designed the methodology. T.L., D.S., and Y.I. acquired the data. T.L. performed data analysis and wrote the initial draft of the manuscript. All authors interpreted the results, revised the manuscript critically, and approved the final version of the manuscript.

---

**Supplementary Figure 1.**
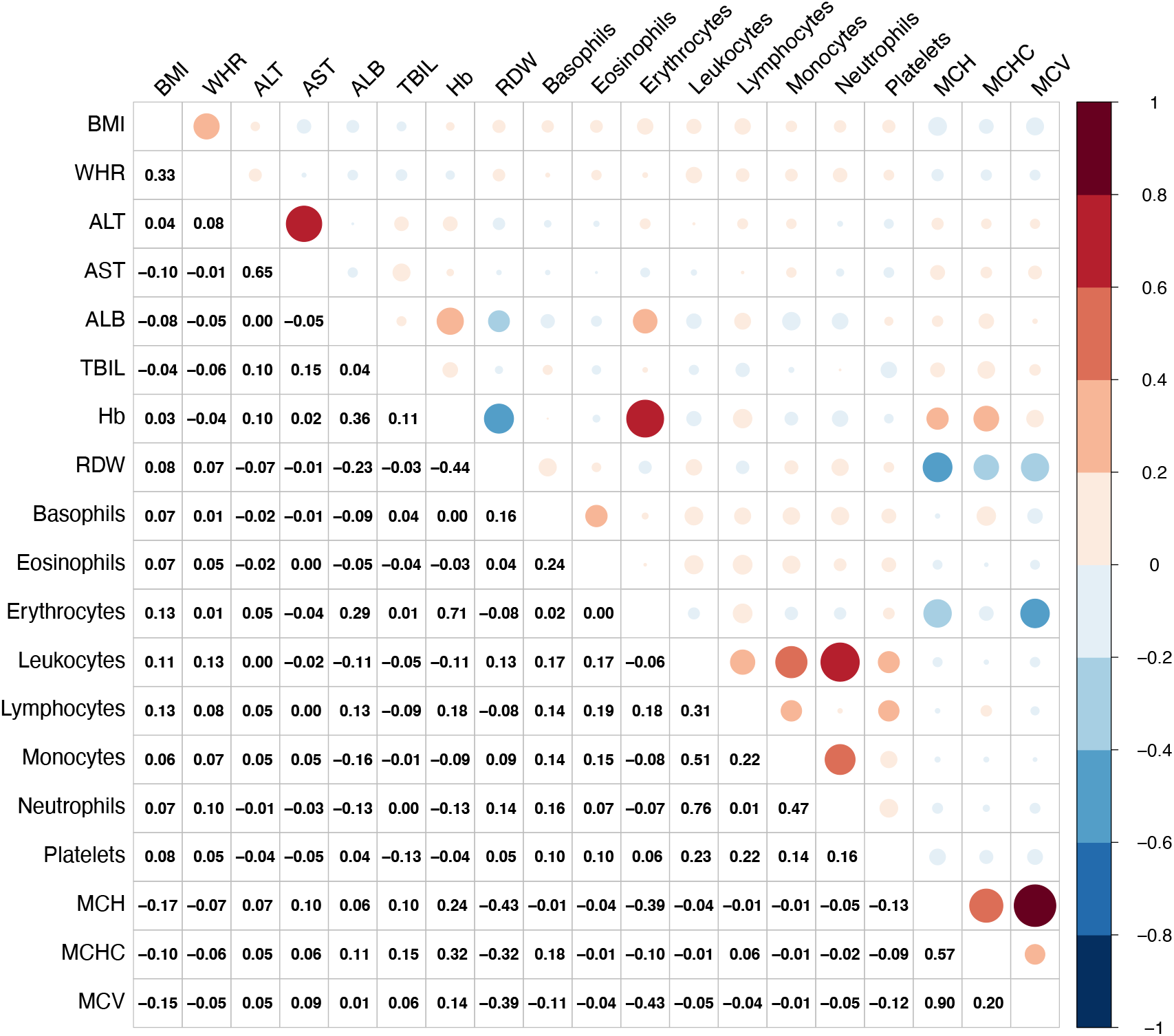
Correlation between candidate phenotypes in the AoU model selection dataset. Pearson correlations were computed using pairwise complete observations. The color of each circle indicates the direction of the correlation (positive or negative), while the size reflects the magnitude. Numerical correlation coefficients are displayed in the lower triangle of the matrix.

## References

1. Cappellini MD, Fiorelli G. Glucose-6-phosphate dehydrogenase deficiency. Lancet Lond Engl. 2008;371(9606):64–74. doi:10.1016/S0140-6736(08)60073-2

2. Nkhoma ET, Poole C, Vannappagari V, Hall SA, Beutler E. The global prevalence of glucose-6-phosphate dehydrogenase deficiency: a systematic review and meta-analysis. Blood Cells Mol Dis. 2009;42(3):267–278. doi:10.1016/j.bcmd.2008.12.005

3. Beutler E. G6PD: Population genetics and clinical manifestations. Blood Rev. 1996;10(1):45–52. doi:10.1016/S0268-960X(96)90019-3

4. Beutler E. G6PD Deficiency. Blood. 1994;84(11):3613–3636. doi:10.1182/blood.V84.11.3613.bloodjournal84113613

5. Hirono A, Kawate K, Honda A, Fujii H, Miwa S. A single mutation 202G>A in the human glucose-6-phosphate dehydrogenase gene (G6PD) can cause acute hemolysis by itself. Blood. 2002;99(4):1498. doi:10.1182/blood.v99.4.1498

6. Jallow M, Teo YY, Small KS, et al. Genome-wide and fine-resolution association analysis of malaria in West Africa. Nat Genet. 2009;41(6):657–665. doi:10.1038/ng.388

7. Nóbrega-Pereira S, Fernandez-Marcos PJ, Brioche T, et al. G6PD protects from oxidative damage and improves healthspan in mice. Nat Commun. 2016;7:10894. doi:10.1038/ncomms10894

8. Yoshida A, Stamatoyannopoulos G, Motulsky AG. Negro variant of glucose-6-phosphate dehydrogenase deficiency (A-) in man. Science. 1967;155(3758):97–99. doi:10.1126/science.155.3758.97

9. Yoshida A, Beutler E. Human glucose-6-phosphate dehydrogenase variants: a supplementary tabulation. Ann Hum Genet. 1978;41(3):347–355. doi:10.1111/j.1469-1809.1978.tb01902.x

10. Karczewski KJ, Francioli LC, Tiao G, et al. The mutational constraint spectrum quantified from variation in 141,456 humans. Nature. 2020;581(7809):434–443. doi:10.1038/s41586-020-2308-7

11. da Rocha JEB, Othman H, Tiemessen CT, et al. G6PD distribution in sub-Saharan Africa and potential risks of using chloroquine/hydroxychloroquine based treatments for COVID-19. Pharmacogenomics J. 2021;21(6):649–656. doi:10.1038/s41397-021-00242-8

12. Wheeler E, Leong A, Liu CT, et al. Impact of common genetic determinants of Hemoglobin A1c on type 2 diabetes risk and diagnosis in ancestrally diverse populations: A transethnic genome-wide meta-analysis. PLoS Med. 2017;14(9):e1002383. doi:10.1371/journal.pmed.1002383

13. Moon JY, Louie TL, Jain D, et al. A Genome-Wide Association Study Identifies Blood Disorder-Related Variants Influencing Hemoglobin A1c With Implications for Glycemic Status in U.S. Hispanics/Latinos. Diabetes Care. 2019;42(9):1784–1791. doi:10.2337/dc19-0168

14. Sarnowski C, Leong A, Raffield LM, et al. Impact of Rare and Common Genetic Variants on Diabetes Diagnosis by Hemoglobin A1c in Multi-Ancestry Cohorts: The Trans-Omics for Precision Medicine Program. Am J Hum Genet. 2019;105(4):706–718. doi:10.1016/j.ajhg.2019.08.010

15. Vuckovic D, Bao EL, Akbari P, et al. The Polygenic and Monogenic Basis of Blood Traits and Diseases. Cell. 2020;182(5):1214–1231 e11. doi:10.1016/j.cell.2020.08.008

16. Hu Y, Stilp AM, McHugh CP, et al. Whole-genome sequencing association analysis of quantitative red blood cell phenotypes: The NHLBI TOPMed program. Am J Hum Genet. 2021;108(5):874–893. doi:10.1016/j.ajhg.2021.04.003

17. Eyth E, Zubair M, Naik R. Hemoglobin A1C. In: StatPearls. StatPearls Publishing; 2025. Accessed June 23, 2025. http://www.ncbi.nlm.nih.gov/books/NBK549816/

18. American Diabetes Association Professional Practice Committee. 2. Diagnosis and Classification of Diabetes: Standards of Care in Diabetes-2024. Diabetes Care. 2024;47(Suppl 1):S20–S42. doi:10.2337/dc24-S002

19. Paterson AD. HbA1c for type 2 diabetes diagnosis in Africans and African Americans: Personalized medicine NOW! PLOS Med. 2017;14(9):e1002384. doi:10.1371/journal.pmed.1002384

20. Breeyear JH, Hellwege JN, Schroeder PH, et al. Adaptive selection at G6PD and disparities in diabetes complications. Nat Med. 2024;30(9):2480–2488. doi:10.1038/s41591-024-03089-1

21. Mardis ER. A decade’s perspective on DNA sequencing technology. Nature. 2011;470(7333):198–203. doi:10.1038/nature09796

22. All of Us Research Program Genomics Investigators. Genomic data in the All of Us Research Program. Nature. 2024;627(8003):340–346. doi:10.1038/s41586-023-06957-x

23. NHLBI TOPMed - NHGRI CCDG: The BioMe Biobank at Mount Sinai. Accessed June 23, 2025. https://www.ncbi.nlm.nih.gov/projects/gap/cgi-bin/study.cgi?study_id=phs001644.v3.p2

24. Bycroft C, Freeman C, Petkova D, et al. The UK Biobank resource with deep phenotyping and genomic data. Nature. 2018;562(7726):203–209. doi:10.1038/s41586-018-0579-z

25. Hocking RR. A Biometrics Invited Paper. The Analysis and Selection of Variables in Linear Regression. Biometrics. 1976;32(1):1–49. doi:10.2307/2529336

26. DeLong ER, DeLong DM, Clarke-Pearson DL. Comparing the areas under two or more correlated receiver operating characteristic curves: a nonparametric approach. Biometrics. 1988;44(3):837–845.

27. Kalakonda A, Jenkins BA, John S. Physiology, Bilirubin. In: StatPearls. StatPearls Publishing; 2025. Accessed June 23, 2025. http://www.ncbi.nlm.nih.gov/books/NBK470290/

28. Schoeler T, Speed D, Porcu E, Pirastu N, Pingault JB, Kutalik Z. Participation bias in the UK Biobank distorts genetic associations and downstream analyses. Nat Hum Behav. 2023;7(7):1216–1227. doi:10.1038/s41562-023-01579-9

29. Pirastu N, Cordioli M, Nandakumar P, et al. Genetic analyses identify widespread sex-differential participation bias. Nat Genet. 2021;53(5):663–671. doi:10.1038/s41588-021-00846-7

30. Zhang X, Gregg EW, Williamson DF, et al. A1C level and future risk of diabetes: a systematic review. Diabetes Care. 2010;33(7):1665–1673. doi:10.2337/dc09-1939

31. Cheng P, Neugaard B, Foulis P, Conlin PR. Hemoglobin A1c as a Predictor of Incident Diabetes. Diabetes Care. 2011;34(3):610–615. doi:10.2337/dc10-0625

32. Pradhan AD, Rifai N, Buring JE, Ridker PM. HbA1c Predicts Diabetes but not Cardiovascular Disease in Non-Diabetic Women. Am J Med. 2007;120(8):720–727. doi:10.1016/j.amjmed.2007.03.022

